# Military sexual trauma-related posttraumatic stress disorder service-connection award denial across gender and race

**DOI:** 10.1101/2023.01.10.23284359

**Authors:** Aliya R. Webermann, Mayumi O. Gianoli, Marc I. Rosen, Galina A. Portnoy, Sally G. Haskell, Tessa Runels, Anne C. Black

## Abstract

The current study characterizes a cohort of veteran claims filed with the Veterans Benefits Administration for posttraumatic stress disorder secondary to experiencing military sexual trauma, compares posttraumatic stress disorder service-connection award denial for military sexual trauma-related claims versus combat-related claims, and examines military sexual trauma -related award denial across gender and race. We conducted analyses on a retrospective national cohort of veteran claims submitted and rated between October 2017-May 2022, including 102,404 combat-related claims and 31,803 military sexual trauma-related claims. Descriptive statistics were calculated, logistic regressions assessed denial of service-connection across stressor type and demographics, and odds ratios were calculated as effect sizes. Military sexual trauma-related claims were submitted primarily by White women Army veterans, and had a two times higher odds of being denied (27.6%) than combat claims (18.2%). When controlling for demographics including age, race, and gender, men veterans had a 1.77 times higher odds of having military sexual trauma-related claims denied compared to women veterans (36.6% vs. 25.4%), and Black veterans had a 1.39 times higher odds of having military sexual trauma-related claims denied compared to White veterans (32.4% vs. 25.3%). Three-fourths of military sexual trauma-related claims were awarded in this cohort, however, there were disparities in awarding of claims across gender and race for men and Black veterans, which suggest the possibility of systemic barriers for veterans from underserved backgrounds and/or who may underreport military sexual trauma.

## Introduction

According to the U.S. Department of Defense, rates of military sexual trauma (MST; sexual harassment and sexual assault experienced during military service) have increased annually since 2018, yet only one-fifth of incidents are reported to military officials [1]. Service members and veterans of all genders, races, and backgrounds experience MST, though MST is substantially more likely to be endorsed among women compared to men [2-4].

Findings vary as to whether MST is more likely to be endorsed by White veterans compared to Black and other racial minority veterans [2, 4]. MST is associated with multiple mental and physical health conditions, most notably posttraumatic stress disorder (PTSD), and veterans who experience MST report more severe PTSD symptoms compared to veterans who experience childhood abuse, non-MST sexual assault, and combat [2, 5].

Veterans who develop or experience worsened mental health issues, including PTSD symptoms, following MST may file a claim with the Veterans Benefits Administration (VBA) within the Department of Veterans Affairs (VA) to receive tax-free monetary compensation and covered healthcare benefits [6]. A 2018 report found that VBA had processed approximately 12,000 PTSD claims related to MST annually in the preceding three years [7]. Veterans Service Representatives (VSRs) review claims, military service records, military treatment records, and post-service treatment records for evidence of MST. VSRs look for “markers” to indicate that symptoms (e.g., PTSD) may have developed or worsened as a consequence of the military stressor (i.e., MST). If VSRs find credible evidence of MST and/or markers, a compensation and pension (C&P) examination is ordered. C&P exams for PTSD are mental health evaluations conducted by doctoral-level clinicians to determine if the veteran has a disability due to a diagnosable mental health condition related to military service. VBA adjudicators consult the C&P exam report with other records to render decisions abouts service-connection, which is calculated using the VA Schedule for Rating Disabilities [6].

Reports from governmental and non-profit organizations have brought attention to issues in the MST C&P process and potential disparities in awarding of MST-related PTSD claims compared to non-MST PTSD claims. Yale Law School, Service Women’s Action Network, and American Civil Liberties Union found that the grant rate for MST-related PTSD claims between 2008-2012 was 16.5% to 29.6% less than the grant rate for non-MST PTSD claims (e.g., combat-related claims) [8]. VA’s Office of the Inspector General reported that approximately half of denied MST claims were not properly processed under VBA procedures, including not ordering a C&P exam when one was warranted, not gathering evidence correctly, and using insufficient medical opinions to decide claims [7, 9]. The percentage of awarded MST-related PTSD claims has increased substantially, from 35.6% in 2011 to 72% in 2021, and award rates are now comparable to non-MST-related PTSD claims [10, 11]. However, rates of MST awards for men veterans have lagged behind those of women veterans; in 2018, the MST grant rate for men was 44.7% compared to 57.7.7% for women [10]. MST-related PTSD claim decisions have not been compared across race, but prior work has found Black veterans are more likely to have PTSD claims denied, and receive lower ratings for PTSD service-connections relative to non-Black veterans, especially when psychometrically-validated diagnostic measures are not used [12, 13].

Given evidence of potential disparities in awarding of MST claims and PTSD claims generally, there is a critical need for additional empirical work to examine PTSD claims broadly, and MST-related PTSD claims specifically. Receipt of PTSD benefits is associated with subsequent stable or increased mental healthcare use among veterans [14], emphasizing the necessity of accurate and fair service-connection decisions. To gain further insight into patterns of awarding of MST-related PTSD claims, we used a nationally representative veteran sample to address the following study aims: 1) Compare PTSD service-connection award decisions for MST versus combat claims; 2) characterize a cohort of veterans who filed for MST-related PTSD service connections within a 5-year time period; and 3) examine differences in MST-related PTSD service-connection award decisions across gender and race.

## Method

### Study design and sample

The present study is a secondary analysis of a retrospective cohort of 125,831 veterans who filed initial (new) and/or supplemental (review or appeal) claims for PTSD service-connected disability benefits claims to VBA, and had claims completed and rated by VBA between October 1, 2017 and May 19, 2022. The data were obtained through a request to VBA and included 31,803 MST-related PTSD claims (92.2% initial), 102,404 combat-related PTSD claims (92.1% initial), and 1,364 claims that included MST and combat. PTSD claims not related to MST or combat were not included in this analysis. The present study was approved by the VA Connecticut Healthcare System Institutional Review Board in December 2022. The project was not considered human subjects research and did not require obtaining participant consent. MST claimants were primarily women (79.7%), non-Hispanic White (55.5%), Army veterans (45.4%), non-commissioned officers (65%), pre-9/11 era (62.6%) and averaged 35.77 years old (*SD* = 8.89, range = 19-79). Combat claimants were primarily men (92.3%), non-Hispanic White (69%), Army veterans (68.3%), non-commissioned officers (80.1%), post-9/11 era (72.4%), and averaged 41.08 years old (*SD* = 8.82, range = 19-87).

### Study variables and analyses

The data set included veteran age, gender, race, ethnicity, period of service, military branch, and rank (see Table 1), and claim information including: service-connection award decision (i.e., awarded or denied); explanation for service-connection award decision (e.g., incurred/caused by service); basis for PTSD service-connection (e.g., MST, combat); percent disability rating from 0-100%; and relevant claim dates, including date of VBA claim receipt, date of claim authorization, and date of rating decision. The majority of study variables were categorical, including all claim information except for disability rating (continuous variable from 0-100%), and identity and military service characteristics except for age. The primary outcome was service-connection award decision, a dichotomous variable (awarded or denied).

**Table 1:** Characteristics of Cohort Submitting MST-Related PTSD Claims (*N* = 31,803)

|  | Service-connection<br>awarded | Service-connected<br>denied | Total (% of MST claimant<br>sample) |
| --- | --- | --- | --- |
| Gender |  |  |  |
| Women | 18,794 (74.6%) | 6,399 (25.4%) | 25,193 |
| Men | 4,054 (63.4%) | 2,343 (36.6%) | 6,397 |
| Race/ethnicity |  |  |  |
| White, not of Hispanic origin | 13,130 (74.7%) | 4,438 (25.3%) | 17,568 |
| Black, not of Hispanic origin | 6,625 (67.6%) | 3,176 (32.4%) | 9,801 |
| Asian or Pacific Islander | 1,019 (73.4%) | 370 (26.6%) | 1,389 |
| American Indian/Alaskan<br>Native | 474 (73.4%) | 172 (26.6%) | 646 |
| Unknown | 1,645 (73.9%) | 582 (26.1%) | 2,227 |
| Hispanic origin | 124 (76.5%) | 38 (23.5%) | 162 |
| Age $M$ ($SD$) | 35.09 (8.82) | 37.54 (8.84) | 35.77 (8.89) |
| Service era |  |  |  |
| Pre-9/11 | 14,428 (72.5%) | 5,472 (27.5%) | 19,900 |
| Post-9/11 | 8,587 (72.1%) | 3,316 (27.9%) | 11,903 |
| Highest rank |  |  |  |
| Non-Commissioned Officers | 14,865 (73.4%) | 5,376 (26.6%) | 20,241 |
| Enlisted Personnel | 6,290 (68.6%) | 2,873 (31.4%) | 9,163 |
| Warrant Officers | 122 (83%) | 25 (17%) | 147 |
| Commissioned Officers | 1,270 (79.5%) | 327 (20.5%) | 1,597 |
| Branch |  |  |  |
| Army | 10,344 (71.7%) | 4,080 (28.3%) | 14,424 |
| Navy | 5,685 (70.9%) | 2,333 (29.1%) | 8,018 |
| Air Force | 4,263 (74.8%) | 1,433 (25.2%) | 5,696 |
| Marine Corps | 2,383 (73.8%) | 845 (26.2%) | 3,228 |
| Coast Guard | 325 (78.5%) | 89 (21.5%) | 414 |
| Other (Space Force,<br>Commissioned Corps, etc.) | 14 (63.6%) | 8 (36.4%) | 22 |

Note: All *n*'s do not = 31,803 due to missing data.

Using the dataset of combat-related and MST-related PTSD claimants, we used a binary logistic regression to model the probability of PTSD service-connection award denial as a function of gender, age, race, and stressor type (combat or MST). All predictors were entered simultaneously. Looking specifically at MST claimants, a second binary logistic regression model estimated the probability of MST service-connection award denial by gender, age, and race, entered simultaneously. The data met assumptions for logistic regression, including independence among predictor variables (VIF = 1.021–2.076).

## Results

### Descriptive information on MST and combat-related PTSD claims

Of the 31,803 MST claims submitted and rated, 72.4% (*n* = 23,015) were awarded and 27.6% (*n* = 8,788) denied. Comparatively, of 102,404 combat claims submitted and rated, 81.8% (*n* = 83,754) were awarded and 18.2% (*n* = 18,650) denied. The average MST-related PTSD service-connection award percentage was 60.35% (*SD* = 17.66), while the average combat-related PTSD service-connection rating was 55.27% (*SD* = 17.94). MST claim processing time was on average 4.78 months (*SD* = 4.41, range = 0-39 months) compared to 3.62 months (*SD* = 3.74, range = 0-40 months) for combat claims. The reasons for PTSD claim denial by VBA for both MST and combat claims were, in order: 1) No diagnosis (i.e., does not meet established diagnostic criteria); 2) Not incurred/caused by service (i.e., meets established diagnostic criteria but not related to claimed stressor(s)); 3) Not aggravated by service (i.e., previously met established diagnosed criteria which were not substantially aggravated by claimed stressor(s)); and 4) Other (e.g., claimed stressor(s) did not occur during military service). For both MST and combat-related claims, service-connection denial rates were highest among Black veterans, Navy veterans, and formerly enlisted personnel, but denial rates were higher among men for MST-related claims and women for combat-related claims.

### Differences in awarding for MST vs. combat-related PTSD claims

Controlling for other variables in the model, MST-related PTSD claims had a 2.00 times higher odds of being denied (27.6% denied) compared to combat-related PTSD claims (18.2% denied), χ ^2^ (7) = 1956.41, *p* < .001 [95% CI = 1.94-2.11].

### Differences in awarding for MST claims across gender and race

Controlling for other variables in the model, men claimants had 1.77 times higher odds of MST service-connection denial (36.6% denied) compared to women claimants (25.4% denied), χ ^2^ (6) = 918.01, *p* < .001 [95% CI = 1.66-1.87]. Black claimants had a 1.39 times higher odds of MST service-connection denial (32.4% denied) compared to White claimants (25.3% denied), χ^2^ (6) = 918.01, *p* < .001 [95% CI = 1.31-1.47].

## Discussion

Three-fourths of MST-related claims were awarded in a 5-year period, however, MST-related PTSD claims were significantly more likely to be denied compared to combat-related PTSD claims. This may be due to differences in the process for MST versus combat claims, including that combat-related claims do not require assessing for markers of occurrence or harm per VBA policy whereas MST-related claims typically have this requirement (6, 7, 10). Additionally, men veterans and Black veterans were more likely to be denied MST-related service-connection compared to White veterans and women veterans, respectively. This may be due to lower MST reporting among men service members and veterans, complicating efforts to find MST markers in men. Other possibilities are differences in PTSD symptom severity between men and women, minimization of PTSD in men, and bias among VA providers and adjudicators in substantiating a diagnosis of MST-related PTSD in men. Men are likely than women to report MST due to concerns about others’ perceptions of their masculinity and/or sexuality, professional and personal retaliation, shame and embarrassment, and because MST is often perpetrated via bullying/hazing which makes men less likely to identify incidents as MST (1, 3). The lower rates of MST awards among Black claimants are consistent with Black veterans’ lower likelihoods of being diagnosed with PTSD via C&P exams and being awarded PTSD service-connections compared to White veterans (4, 12).

This study was strengthened through using a large and nationally representative sample of PTSD claims filed, completed, and rated over a five-year period. The study was limited by not including other potentially important covariates impacting MST-related PTSD service-connection awards such as PTSD symptom severity, comorbid mental and physical health issues and/or diagnoses (e.g., depression, substance use disorders), and additional claims submitted other than combat and MST-related PTSD claims. Furthermore, our sample only included binary gender, likely based on sex assigned at birth (i.e., male or female), and was not inclusive of identity characteristics outside of race, gender, and age, such as sexual orientation. Future research may consider including symptom severity measures as a potential covariate, and examining claim re-ratings over time. Additionally, more work is needed to examine diverse veteran experiences with and perspectives on the MST claims process and how those intersect with veteran gender, race, and other identity characteristics. The findings of this study suggest the possible value of anti-bias training for VBA and VHA staff who work within the MST claims process, and programs to encourage active duty personnel and veteran reporting of MST and MST-related PTSD claim submissions, especially for men and Black veterans.

## Data Availability

Given restrictions imposed by good ethical practice and requirements of the U.S. Department of Veterans Affairs Institutional Review Board regarding protection of human subjects data, and consistent with journal guidelines for cases where there are ethical restrictions to uploading the data publicly, such as the data being potentially identifying or a lack of patient consent to publicly share the data, we will make the data available to qualified researchers upon request. We have identified a non-author contact point who will process all data requests and provide access to de-identified data as appropriate. The non-author contact is Praveen Kamath and he can be contacted at.

## Acknowledgements

None.

## Notes

The opinions expressed in this article are those of the authors and do not represent the official policy or position of the U.S. Department of Veterans Affairs or the U.S. government. Dr. Webermann’s time is funded by the Advanced Fellowship in Women’s Health through the VA Office of Academic Affiliations. The funding sources had no involvement in the study design, analyses, article preparation, or decision to submit. The authors declare no conflicts of interest.

### Competing Interest Statement

The authors have declared no competing interest.

### Funding Statement

The author(s) received no specific funding for this work. Dr. Webermann’s time is funded by the Advanced Fellowship in Women’s Health through the VA Office of Academic Affiliations. The funding sources had no involvement in the study design, analyses, article preparation, or decision to submit.

### Author Declarations

The present study was approved by the VA Connecticut Healthcare System Institutional Review Board in December 2022

